# Self-medication practices for oral health problems: A community-based cross-sectional study in Sri Lanka

**DOI:** 10.1101/2024.11.29.24318207

**Authors:** Kavithrini Anunadika Gammulle, Sudeha M Premarathne

## Abstract

The prevalence of self-medication is a common practice among Sri Lankans. However, the practice of self-medication for oral health problems has not been studied in the country. The current study aimed to assess the prevalence and the associated factors of self-medication practices for oral conditions. This community-based cross-sectional study was conducted among 441 adults in Piliyandala, Sri Lanka. The participants who had experienced an oral health-related concern within the past year were selected using a multi-stage cluster sampling method with probability proportionate to size technique. An interviewer-administered questionnaire was used for data collection. The results revealed that 68.9% practiced self-medication for an oral health condition during the past year. Dental pain was the most common reason for self-medicating. Allopathic medications were used by 85.2%. Amoxicillin and Metronidazole were taken by 10.2% and 12.8% respectively. Poor attitude towards oral healthcare services and low family income were significant predictors of self-medication and despite the availability of oral health services, a considerable number of participants resorted to self-medication due to the perceived simplicity of the condition and the high cost of dental treatments.

## Introduction

Self-medication is the use of medicinal products by individuals to treat self-recognized disorders or symptoms without professional guidance (1). Despite the availability of free health care, individuals in Sri Lanka resort to self-medication to address general disorders (2), without proper professional guidance. This trend raises concerns regarding the potential risks associated with inadequate treatment practices, including the delay in diagnoses, the emergence of antimicrobial resistance, and the possibility of new health conditions due to improper medication use. This phenomenon is alarming given that self-medication behaviors can rely on a combination of cultural and socio-economic factors (3). Although there are a few benefits of self-medication such as increased treatment options for minor ailments, enhanced access to medicines, empowering individuals, and less burden on the healthcare system, the potent health risks including incorrect diagnoses, contraindicated medication use, increased resistance to antimicrobials, adverse drug reactions, and delayed professional care, may outweigh the advantages(4). The high prevalence among the elderly (5), the pregnant (6,7), and breastfeeding mothers (8) cannot be disregarded. Antimicrobial resistance itself is estimated to result in an annual economic loss of US$ 100 trillion and 10 million deaths by 2050. Therefore, WHO urged member countries to develop national strategies to mitigate this public health issue(9). Nevertheless, over half of the population in Asian countries self-medicate (10–12).

The popularity of self-medication in developing countries highlights the urgent need for public health interventions to educate individuals on the risks related to incorrect self-diagnosis and the inappropriate use of medications(13). In Sri Lanka, the prevalence of self-medication with antibiotics ranged from 2.6% in rural (14) to 11% in urban settings (15). Self-medication with allopathic, traditional, and home remedies was found to be 33.9%, and 35.3% in urban and rural regions in that order, and relied upon the satisfaction with health services and the likelihood of visiting medical services(2). However, the prevalence of self-medication practices for oral health issues in Sri Lanka is not well-documented. Throughout the years, the prevalence of self-medication has increased with the expansion of the pharmaceutical industry, lack of regulation by the responsible authorities(16) and the COVID-19 pandemic (17,18).

However, self-medication for oral conditions is unexplored in Sri Lanka. Therefore, bridging this gap, the current study assessed the prevalence and determinants of self-medication for oral health problems among Sri Lankan adults, and how this practice relates to dental healthcare services.

## Methods

### Study design, setting, sample size, and sampling method

This study was a community-based cross-sectional study conducted in 15 Public Health Midwife (PHM) areas in the Piliyandala Medical Officer of Health (MOH) area in Colombo, Sri Lanka. Every participant was provided with information on the study and informed written consent was obtained before engaging them in the study. The period of data collection spanned from 1^st^ August to 30^th^ November 2019. It was carried out among people who have had an oral health-related concern within the past year and are 18 to 60 years old.

The sample size was calculated using the formula for estimating a population proportion with absolute precision (19). There were no studies on the prevalence of self-medication for oral conditions among Sri Lankans. An Indian study revealed a 51.63% prevalence of self-medication for oral conditions (20) and it was considered the estimated prevalence of self-medication. With a 95% confidence level, a minimum sample size of 385 was required. However, after incorporating a design effect of 1.1, and a 5% non-responsive rate, the final sample size was 444.

Further, multistage cluster sampling with the Probability Proportionate to Size Sampling (PPSS) technique was employed to select the sample. In the first stage, PHM areas in the Piliyandala MOH area were considered cluster units. It was decided to include 30 subjects in a cluster. As the calculated minimal sample size was 444, the number of clusters required was 15. Therefore, it was decided to include 15 clusters and a sample size of 450. A random number that is less than the sampling interval was generated by using random number-generating software. The random number was 3520. The first cluster was selected from the PHM area that had the 3520th person in the cumulative population. Then the sampling interval of 6720 was added to the random number. It gave the value of 10240. The second cluster was selected from the PHM area which had the value of 10240 in the cumulative total. The remaining clusters were selected using the same method and each time, the sampling interval was added to the previously selected number.

In the second stage, from each selected PHM area, one road was randomly selected. The required sample size from the particular PHM area was collected from the residents who lived on both sides of that road. The first house was randomly selected from the eligible family register (a register that the PHMs maintain) followed by the house to the left of the first house as the second house to be selected. The rest of the houses were chosen by employing the same method. Data collection along one road was carried out until it fulfilled the cluster size. The closest road to the left of the selected road was screened if the required number of study units couldn’t be found from the selected road.

### Ethics Statement

Ethical approval was obtained from the Ethics Review Committee, Postgraduate Institute of Medicine, University of Colombo (ERC/PGIM/2019/106). All procedures performed in the study followed the ethical standards of the institute and the 1964 Helsinki Declaration and its later amendments.

### Data Collection Method

An interviewer-administered questionnaire contained both close-ended questions and open-ended questions. The first section assessed socio-demographic factors, dental service utilization, accessibility to dental services, and knowledge of complications of self-medication. Section two inquired about the nature of care sought for oral health problems, history of self-medication, factors related to self-medication, and opinion on dental services. The questionnaire was pre-tested with 25 individuals from the Kesbewa MOH area and modified to improve the face and content validity.

Data was collected during weekends and weekdays, mornings, and evenings to include both working and non-working populations. PHMs were trained to administer the questionnaire alongside the PI. Data collection in adjacent PHM areas was done simultaneously on the same day to reduce cross-contamination. Following data collection, a printout that consisted of information on available government dental services in the area was distributed among the participants.

Data was then entered into an Excel spreadsheet and imported to R version 4.4.0 (2024-04-24 ucrt). Before data analysis, a rigorous data cleaning process was carried out to account for the missing values and outliers to minimize biases and inaccuracies. Data analysis was initiated with descriptive statistics that summarized the demographic and socio-economic characteristics of the participants, the prevalence of self-medication, the types of medications, and reasons for self-medication. Following this, univariate analysis between the independent variables (gender, age, level of education, employment status, family income, distance to the nearest government dental clinic, distance to the nearest private dental clinic, last visit to a dental clinic, opinion regarding dental services, and knowledge of adverse reactions) and the dependent variable (the practice of self-medication) was carried out using the Chi-square test and Fisher’s exact test (when the expected cell count was less than 5). Multiple logistic regression was then performed to identify the predictors of self-medication practices. The analysis included step-wise backward selection which had a better Akaike Information Criterion (AIC) than the full model.

## Results

### Demographics

The study achieved a response rate of 98% which consisted of 441 participants. Females constituted 58.6% of the sample and the majority (63%) were less than 40 years. A significant proportion of the participants had not continued education beyond school (86.1%) and approximately half (45.6%) were unemployed. Most (65%) participants had a monthly income of less than 50,000 Sri Lankan rupees.

### Self-medication Practices

The prevalence of self-medication was 68.9% in this sample. Dental pain was the most common oral health problem that warranted self-medication, reported by 67.1% of participants. Among those who did not self-medicate, 71.5% consulted a dental practitioner, 24.7% sought care from a medical doctor, and 3.6% received treatment from an Ayurvedic doctor. Allopathic medicines were used by 85.2% of those who self-medicated and paracetamol was the most frequently used medication (45.5%), followed by non-steroidal anti-inflammatory drugs (NSAIDs) (31.5%). Further, a smaller percentage of 10.2% and 12.8% in that order have used Amoxycillin and Metronidazole without prescription (Figure.1). Most participants (64.8%) obtained the medicines for self-medication from pharmacies. A significant proportion of the participants (67.4%) reported resorting to self-medication because they perceived their oral health problem as minor. High treatment costs for dental diseases motivated 38% of them to self-medicate, while 3% did not seek professional care because they lacked confidence in dentists. Among the study participants, awareness of the adverse effects and complications of self-medication was relatively high, and 25.6% of them were aware of antimicrobial resistance.

**Figure 1.**
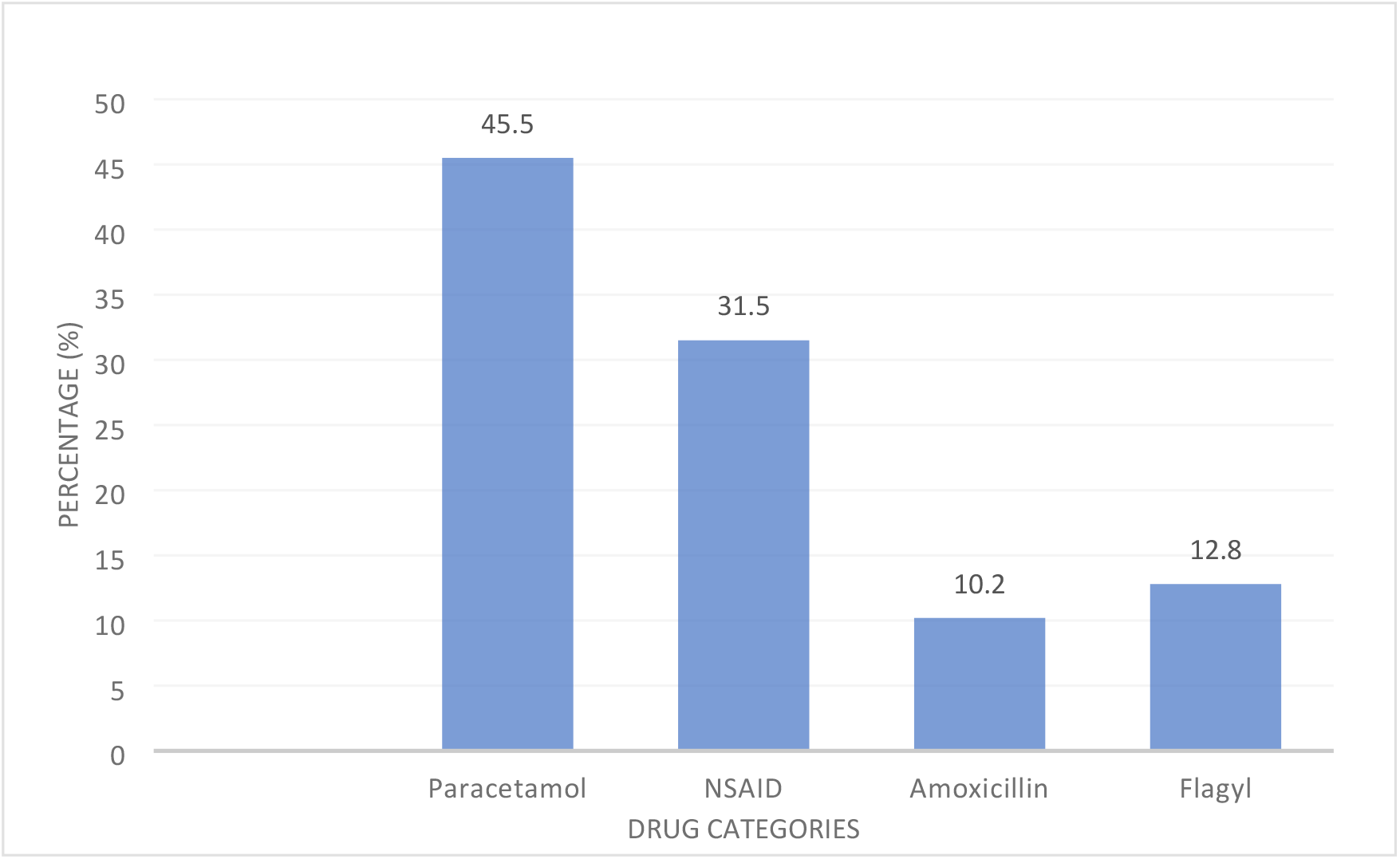
Percentage Usage of the Drug Categories

### Access to and opinion about dental services

Over half of the participants (59%) lived within 1km of a government or privately-owned dental clinic, and only 5% lived more than 8km away. However, about 6% had never visited a dentist, and over 40% had not visited one in more than a year. Importantly, about 12% of them were not satisfied with the quality of services they received at the government and private dental clinics they visited.

### Univariate analysis

Univariate analysis revealed that the associations between the practice of self-medication with knowledge of adverse effects, level of education, employment status, and family income were statistically significant (Table 1).

**Table 1.** Univariate Analysis of Variables Associated with Self-Medication Practices among the Population in Piliyandala, Sri Lanka, 2019.

| Variable | Chi square | Significance |
| --- | --- | --- |
| Gender | 0.01 | 0.92 |
| Age | 0.02 | 0.89 |
| Level of education | 9.81 | <0.05 |
| Employment status | 15.25 | <0.05 |
| Family Income | 89.32 | <0.05 |
| Distance to a government clinic | 0.52 | 0.47 |
| Distance to a private clinic | 0.64 | 0.42 |
| Last visit to a dental clinic | 3.48 | 0.06 |
| Opinion regarding dental services | 3.29 | 0.07 |
| Knowledge of adverse reactions | 9.21 | <0.05 |

*Note: The degree of freedom (df) for all Chi-square tests is 1.*

### Multiple logistic regression

Predictors that showed significance in the univariate analysis and those that significantly impacted self-medication according to literature were retained in the initial model of multiple logistic regression. Stepwise backward selection improved the Akaike Information Criterion compared to the full model. The final model however included two predictors namely opinion regarding dental services and family income. Both predictors were statistically significant, with family income having a particularly strong prediction capability toward self-medication (Table 2).

**Table 2.**
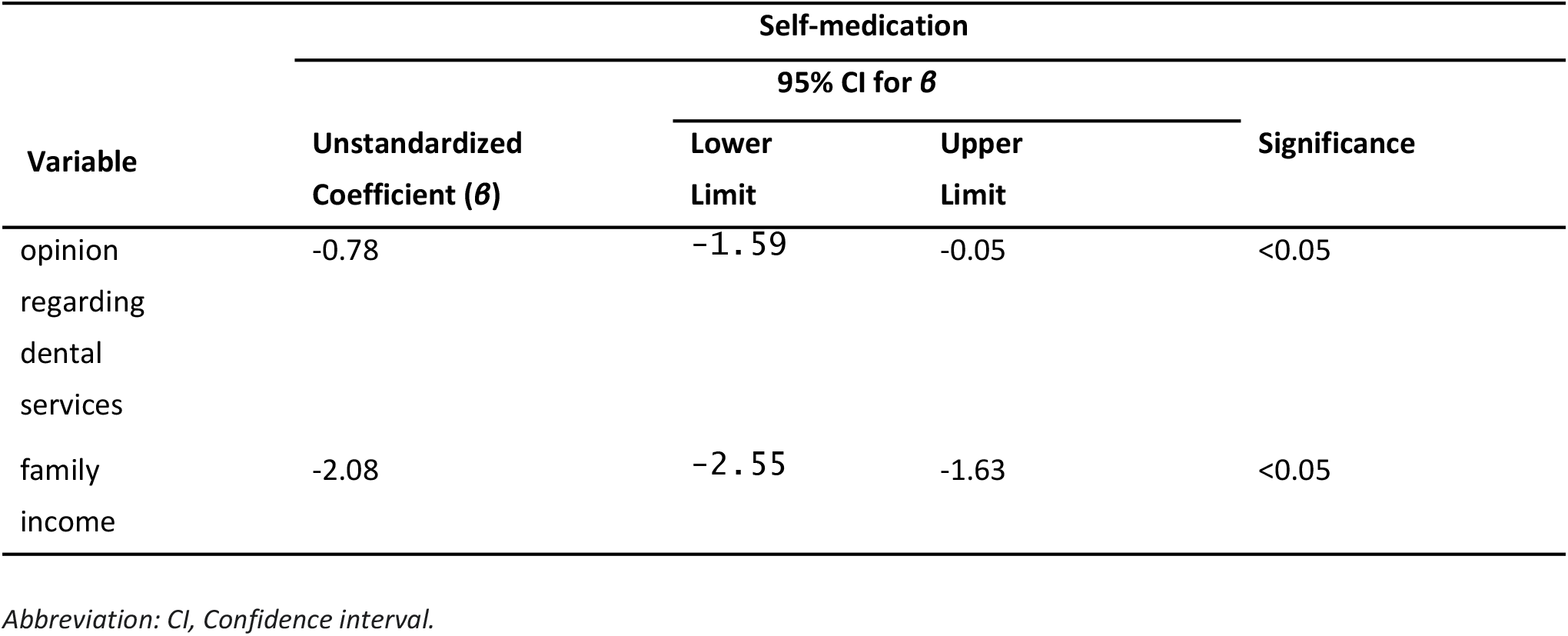
Multiple Logistic Regression Analysis for Predictors of Self-Medication Practices among the Population in Piliyandala, Sri Lanka, 2019.

## Discussion

This study explored the practice of self-medication for oral diseases in Sri Lanka. The participants were mostly females aged less than 40 years and about half of them were unemployed with a monthly income of less than 50,000 Sri Lankan rupees. The study revealed that 68.9% of the study participants have self-medicated for oral disease, a higher prevalence compared to some studies (21) and similar to some (22). However, the prevalence of self-medication for other systemic disorders is found to be lesser in Sri Lanka (2) and other countries (23,24)

Antibiotics were consumed by 10-13% of our study which was comparable to findings from previous Sri Lankan studies on self-medication for general disorders (15). The majority of the participants had obtained the medicines from a pharmacy, without a prescription. According to previous literature, this has been a common practice in Sri Lanka (25). Despite the availability of free dental services, the high cost of dental treatments in the private sector has directed 38% of them to self-medicate. It is beyond the scope of this study to assess the factors that draw patients to the private sector whilst the government sector dental clinics function most days of the week. The results suggest that the practice of self-medication significantly relies upon the consumer’s opinion towards oral healthcare services and is more common among those who belong to the lower socio-economic strata.

Therefore, this research underscores that it is necessary to improve awareness regarding responsible self-medication, targeting those who belong to the lower socio-economic strata. It also highlights that the attitude towards oral health service provision is required to be positive for more effective utilization of services.

A limitation of the study was the potential sampling bias that could arise from the overrepresentation of the unemployed and the retired in the population due to data being collected from households. Further, response bias due to the underreporting of self-medication and recall bias may have introduced confounding despite stringent efforts to mitigate the biases.

The authors are confident that the current study provides strong evidence to inform policy decisions to strengthen the utilization of healthcare services and address the socioeconomic disparities of access to the services. Future studies may explore how self-medication can be manipulated and navigated to ease the burden on the free delivery of healthcare services in Sri Lanka.

## Conclusion

This study highlights the problem of self-medication for oral diseases in Sri Lanka despite free oral healthcare delivery. The results suggest that fostering positive attitudes towards oral healthcare services while addressing socioeconomic disparities is crucial for reducing self-medication practices for oral health problems in Sri Lanka.

## Data Availability

All relevant data is within the manuscript and its supporting information files.

